# Tuberculosis and COVID-19 in 2020: lessons from the past viral outbreaks and possible future outcomes

**DOI:** 10.1101/2020.04.28.20082917

**Authors:** Radu Crisan-Dabija, Cristina Grigorescu, Cristina-Alice Pavel, Bogdan Artene, Iolanda Valentina Popa, Andrei Cernomaz, Alexandru Burlacu

**Affiliations:** University of Medicine and Pharmacy “Grigore T. Popa” Iasi, Pulmonology Department, Head of Clinic of Pulmonary Diseases Iasi; University of Medicine and Pharmacy “Grigore T. Popa” Iasi, Depatment Thoracic Surgery, Clinic of Thoracic Surgery Iasi, Hospital of Pulmonary Diseases Iasi; Clinic of Pulmonary Diseases Iasi, Romania; Department of Interventional Cardiology - Cardiovascular Diseases Institute, Iasi, Romania; Institute of Gastroenterology and Hepatology, Iasi, Romania, and ‘Grigore T. Popa’ University of Medicine, Iasi, Romania; University of Medicine and Pharmacy “Grigore T. Popa” Iasi, Pulmonology Department, Institute of Oncology Iasi; Head of Department of Interventional Cardiology Cardiovascular Diseases Institute, and “Grigore T. Popa” University of Medicine, Iasi, Romania

**Keywords:** SARS-CoV-2, COVID-19, tuberculosis, SARS, coinfection, MERS

## Abstract

**Background:** The threat of contagious infectious diseases is constantly evolving, as demographic explosion, travel globalization and *changes in human lifestyle increase the risk* of spreading pathogens, leading to accelerated changes in disease landscape. Of particular interest is the aftermath of superimposing viral epidemics (especially SARS-CoV-2) over long-standing diseases, such as tuberculosis (TB), which remains a significant disease for public health worldwide and especially in emerging economies.

**Methods and Results:** PubMed electronic database was requested for relevant articles linking TB, influenza and SARS-CoV viruses and subsequently assessed eligibility according to inclusion criteria. Using a data mining approach, we also queried the COVID-19 Open Research Dataset (CORD-19). We aimed to answer the following questions: What can be learned from other coronavirus outbreaks (with a focus on TB patients)? Is coinfection (TB and SARS-CoV-2) more severe? Is there a vaccine for SARS-CoV-2? How does the TB vaccine affect COVID19? How does one diagnosis affect the other?

**Discussions:** Few essential elements about TB and SARS-CoV coinfections were discussed. First, lessons from the past outbreaks (other coronaviruses), as well as influenza pandemic / seasonal outbreaks have taught the importance of infection control to avoid the severe impact on TB patients. Second, although challenging due to data scarcity, investigating the pathological pathways linking TB and SARS-CoV-2 leads to the idea that their coexistence might yield a more severe clinical evolution. Finally, we addressed the issues of vaccination and diagnostic reliability in the context of coinfection.

**Conclusions:** Because viral respiratory infections and TB impede the host’s immune responses, it can be assumed that their harmful synergism may contribute to more severe clinical evolution. Despite the rapidly growing number of cases, the data needed to predict the impact of the COVID-19 pandemic on patients with latent TB and TB sequelae still lies ahead.

## 1. Introduction

The global threat of contagious infectious diseases – in particular, tuberculosis (TB) – has long concerned authorities in charge with public health policies. Most data and all predictions concerning global epidemiology of TB are based upon “*real-life*” analysis (surveys and national surveillance programmes) conducted by the World Health Organisation (WHO) (1, 2). The incidence of TB is slowly declining, but still remains an important issue worldwide (ranked as the ninth leading cause of death worldwide and the leading cause from a single infectious agent (3, 4)), especially in most middle-income and emerging-economy countries.

TB remains of great significance for the public health in Eastern Europe (e.g. Romania), which has the highest TB incidence in the European Union (EU) (4 times higher the average), accounting for a quarter of the TB burden in the EU (4). The incidence of TB increased in Romania after 1990, peaking in 2002 (142.2%), with a downward trend since then, reaching 54.5/100 000 in 2016, 54.2% lower than in 2002 (4, 5). A series of factors augmented the severity of TB endemic in Romania, namely: a large number of severe forms, cases with multidrug-resistant tuberculosis (MDR-TB) and extensively drug-resistant TB (XDR-TB), HIV coinfection, and (TB-related) mortality in children. TB mortality in Romania has followed the same course as the incidence, with a peak in 2002 and an elevation of XDR-TB cases between 2012 and 2015, with a three-fold increase (4).

Influenza infection may promote the progression of latent Mycobacterium tuberculosis (MTB) infection to active TB, alter the clinical presentation of TB, and also possibly exacerbate pulmonary TB (PTB) (6). Both influenza and tuberculosis hinder host immune responses. Specifically, influenza can impair T cell immunity and weaken innate immune responses against secondary bacterial infections (6, 7).

This deleterious synergism of viral and bacterial infections increases the risk of influenza associated mortality, and for patients with PTB may increase the severity of influenza disease and of death due to chronic lung disease and immunosuppression. Epidemiologic data suggest an increased rate of influenza disease or severe influenza associated disease in patients with TB during influenza pandemics (6, 8, 9) or during seasonal influenza epidemics (10) compared with non-TB individuals.

## 2. Objectives

Individuals with chronic respiratory infections, including TB, are first to experience the adverse effects of a pneumotropic pandemic, especially in the healthcare setting (11, 12). Given that both coronavirus disease 2019 (COVID-19) and TB become important causes of mortality worldwide (3, 6, 12) and the TB endemic situation in Romania (4), we sought to explore the possible outcomes of the inevitable collide of the two pandemics. Considering SARS-CoV-2 high transmissibility, it is very likely that COVID-19 will be of particular concern for individuals infected with MTB. (13, 14). Also, coinfection with MTB is of particular importance as the TB diagnosis might be missed or shadowed by concern about COVID-19.

Therefore, we aimed to review the available literature in order to: a) predict the impact of COVID-19 pandemic on patients with latent TB and TB sequelae based on the data available from the past influenza pandemic and seasonal influenza outbreaks (considering similar or more severe outcomes in the current pandemic); b) underline possible clinical particularities and diagnostic errors on these patients; c) evaluate possible different therapeutic approaches on TB patients (latent, sequelae or active) given that current COVID-19 treatment may induce mycobacterial proliferation (15).

## 3. Methods

The electronic database of PubMed was systematically searched for relevant articles from the inception until March 2020. The search terms used were [“*tuberculosis*” OR “TB”], AND [“*flu*” OR “*influenza*”], AND [“*SARS*” OR “*SARS-CoV*” OR “*SARS-CoV-1*”], AND *MERS-CoV*. The search process included article identification, removing the duplicates, screening titles and abstracts, and assessing eligibility of the selected full texts. Additionally, reference lists of valid articles were checked for studies of relevance. Articles were included if they involved data about past TB, SARS-CoV-1, MERS-CoV epidemics, TB - Influenza viruses and TB - SARS-CoV-1 coinfections or clinical or laboratory research on the immune responses during coinfections. Journal articles published with full text or abstracts in English were eligible for inclusion.

In order to identify emerging coinfection particularities of novel coronavirus SARS-CoV-1 we queried the COVID-19 Open Research Dataset (CORD-19), the current largest open dataset available with over 47000 scholarly articles, including over 36000 with full text about COVID-19, SARS-CoV-2 and other coronaviruses from the following sources: PubMed’s PMC open access corpus, a corpus maintained by the WHO, bioRxiv and medRxiv preprints. The CORD-19 dataset is available at https://pages.semanticscholar.org/coronavirus-research. Given the large quantity of textual data in CORD-19, we applied a data mining approach to answer few questions: 1. What can one learn from other coronaviruses epidemics (with a focus on TB patients)? 2. Is coinfection (TB & SARS-CoV-2) more severe? 3. Is there a vaccine for SARS-CoV-2? How does the TB vaccine influence COVID-19? 4. How does one condition influence diagnosis of the other one?

Articles were exported from CORD-19 and merged locally for further processing. Articles of interest were retrieved by administering the query "COVID" OR "COVID-19" OR “2019-nCoV” OR “SARS-CoV-2” OR “Novel coronavirus” OR “Tuberculosis” OR “Mycobacterium tuberculosis” OR "Flu” OR "Influenza” OR “Coinfection” OR “Vaccine” OR „Immunization”. Data mining was further applied in order to select only articles that met our topics of interest about coinfections between particular pathogens stated earlier and COVID-19 developing vaccines. Study selection process and number of papers identified in each phase are illustrated in the flowchart (Figure 1).

**Figure 1 Legend.**
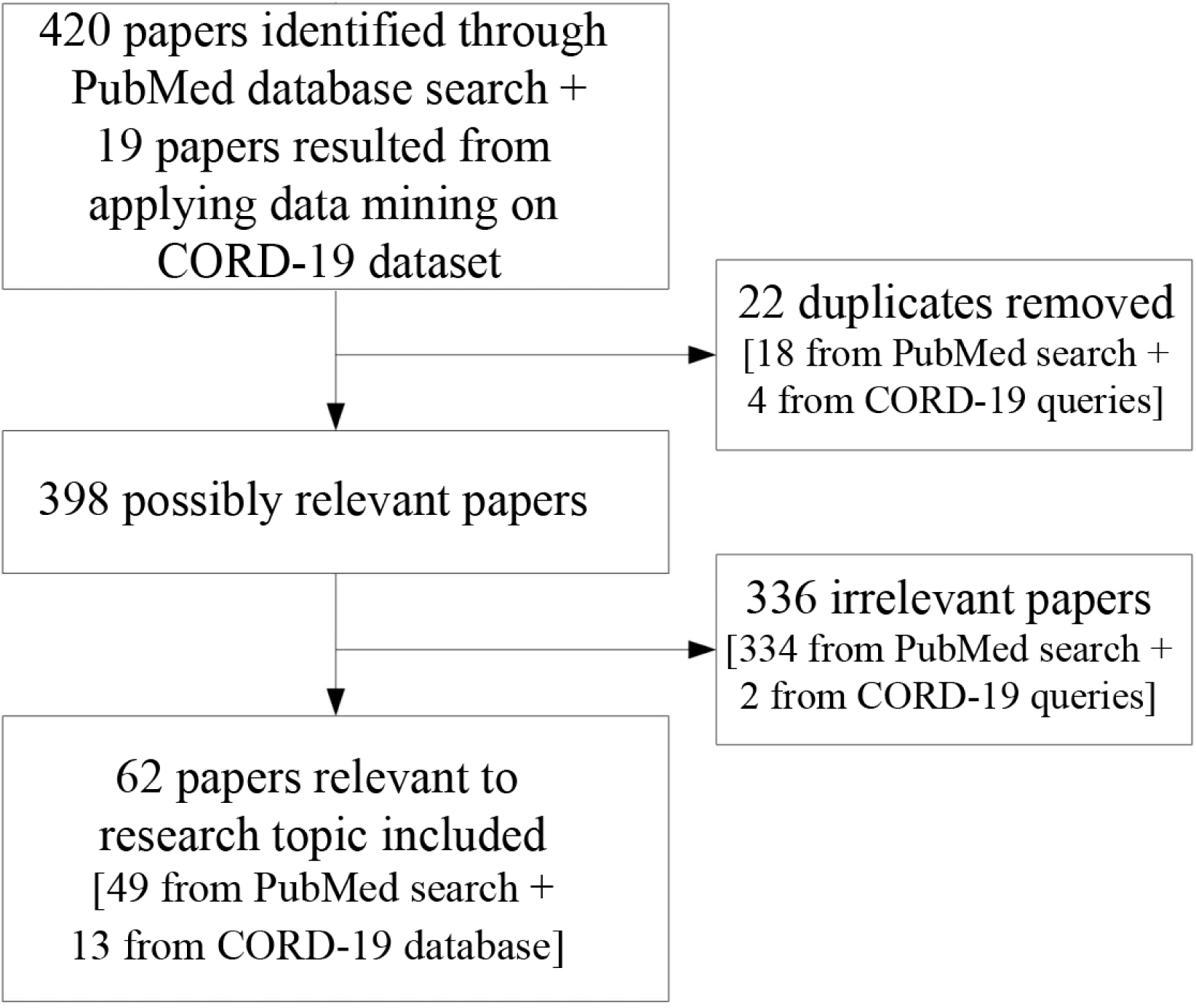
Study selection process and number of papers included.

## 4. Discussions

### 4.1. What can one learn from other past epidemics/pandemics?

For a better understanding of managing a novel coronavirus pandemic, one needs to understand the past experience. Since the first discovery of coronaviruses in 1960, there have been described three human coronaviruses known to cause fatal respiratory diseases: a) the severe acute respiratory syndrome (SARS) coronavirus (SARS-CoV, now known as SARS-CoV-1) that led to a global epidemic in 2002 (13); b) the Middle-East respiratory syndrome coronavirus (MERS-CoV) which was discovered in 2012 and still affects people from 27 countries (16), and most recently, c) the novel coronavirus (SARS-CoV-2), whom outbreak led to an ongoing pandemic with thousands of new cases being confirmed each day and a growing number of reported deaths worldwide (13) (Figure 2).

**Figure 2 Legend.**
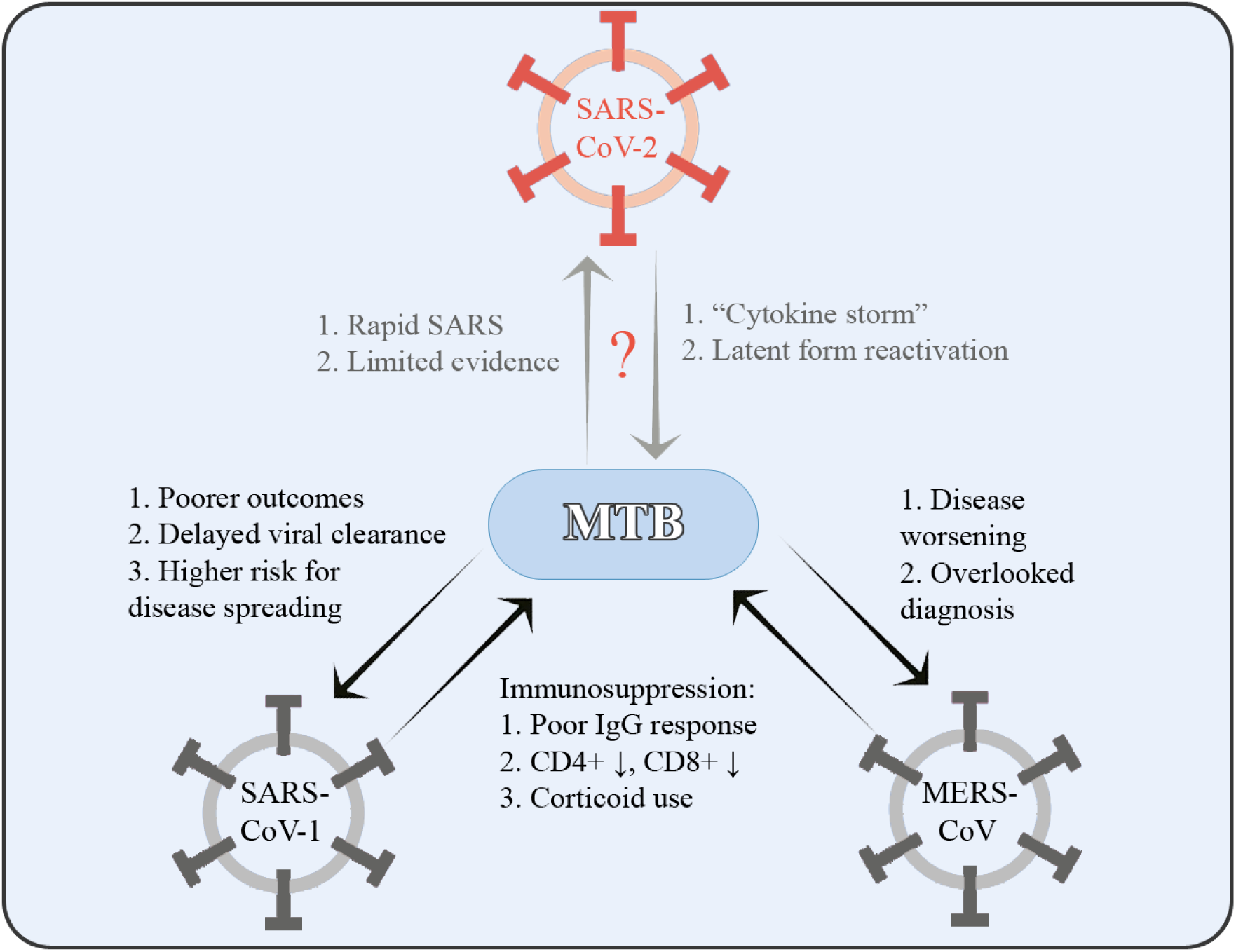
Known and possible interactions between MTB and coronaviruses.

It has to be added that while SARS-CoV-1 was associated in 37 countries with 8096 cases and 774 deaths during the entire 9 months of the epidemic (13) and MERS with only 2494 cases and 858 deaths in 27 countries (13), SARS-CoV-2 spread (and still spreading) in 208 countries with 1009625 confirmed COVID-19 cases and 51737 confirmed deaths (as of the 3^rd^ of April 2020) in only 3 months since the first declared case of COVID-19 pneumonia (17).

Its high transmissibility rate reminds of the 1918–19 influenza pandemic when it’s been estimated almost third of the world’s population being affected with a mortality rate of 2.5% (9). Other significant differences between influenza pandemics and current novel coronavirus pandemic can be found. One of the most notable is that death was less frequent amidst healthcare workers in influenza pandemics as it was the case in SARS, MERS and now COVID-19 pandemic (18). Despite this, other similarities still exist between SARSCoV-2 and influenza as like the striking resemblance of pathological features documented in COVID-19 associated acute respiratory distress syndrome (ARDS) and H7N9-induced ARDS (19). Also, it has been suggested that influenza viruses as well as SARS-CoV-2 significantly upregulate angiotensin-converting enzyme 2 (ACE2) receptors (20). This facilitates novel coronavirus entrance into host cell and makes patients infected by influenza more susceptible to SARS-CoV-2.

It has been shown that in a patient with TB, induction of type I interferons (IFNs) determined by influenza infection could be detrimental (11), impeding the immune competent host’s ability to limit bacterial replication thus promoting the infection (21) and precipitating TB mortality rate (pneumonia and influenza death rates amongst the age group most affected by TB exceeded in 1918 the TB mortality rate noted before and after the pandemic) (9). Also, higher TB death rates were noted in winter months (coinciding with seasonal influenza outbreaks) which led to the suggestion of PTB being an independent risk factor for influenza-associated mortality (10).

During the 2002–03 SARS-CoV-1 epidemic it was highlighted that in order to contain the epidemic, the correct management of symptomatic patients (within and outside hospital) was critical (22). The secondary transmission within Vancouver (Canada) was stopped due to the correct management of several imported cases, as opposed to other places (e.g. Toronto, Canada or Taipei, Taiwan) where the incorrect management conducted to further spread and hospital clusters (22, 23). Also, inappropriate implementation of infection control strategies in Singapore led to massive healthcare personnel infection (half of the SARS cases were among healthcare workers) and several super-spreading events (23).

TB in SARS patients has been reported in several studies from TB endemic countries such as Singapore, China or Taiwan (24, 25), all with known TB patients that acquired SARS and in individuals that developed TB after recovery from SARS (25). The transiently immunosuppression characterized both conditions (26), a reason for poorer IgG antibody response and a delayed viral clearance in coinfected SARS patients (24). Also, use of corticoid therapy in SARS added even more on immunosuppression (24).

During an epidemic, many measures are taken (especially in hospitals) to limit the transmission of the disease to naïve patients. However, overcrowding hospitals is prone to mistakes. Known-TB patients from China, supposedly acquired SARS due to exposure to SARS patients from the same hospital wards, hence coinfection could have been avoided (24). Even though most of them recovered without complications, SARS coinfection on TB cases led to significantly lower mean CD4+ and CD8+ T cells and to undetectable or unusually low antibody levels after SARS recovery (24). Also, the viral excretion was two times longer in sputum and five times longer in stools for TB + SARS patients compared to SARS patients without TB, which translates into a higher potential to spread the virus (24).

When dealing with a possible SARS patient from an endemic TB region, one should never forget TB as a coexisting pathology. In April 2003, a SARS-related hospital screening from Taipei (Taiwan) resulted in discovering 60 TB cases among health-care workers (27) Moreover, during the SARS-CoV-1 epidemic from Singapore, SARS cases were reported developing active PTB short after recovering from SARS (25), data compatible with previous studies on mice regarding the suppression of cellular immunity after a viral infection (11). There is also data on MERS-CoV augmenting TB by the added immunosuppression and reinforcing the need of carefully evaluate a suspected patient (28).

#### Keypoints

1. Influenza pandemic/seasonal outbreaks and other coronaviruses epidemics have a negative impact on TB patients.
2. Transmission prevention was crucial for containing the epidemics.
3. In order to decrease the opportunity of SARS-CoV-2 spreading amongst TB cases, hospital treatment for TB patients should be limited to severe cases.

### 4.2. Is coinfection more severe? Pathological pathways linking TB and SARS-CoV-2

Although the pathophysiology of SARS-CoV-2 is not fully understood, it seems most likely similar to the one of SARS-CoV-1. Solid evidence suggest that SARS-CoV-2 infection could initiate an aggressive inflammation by increasing cytokines secretion such as interleukin-1β (IL-1β), interferon-*γ* (IFN-*γ*), tumour necrosis factor-α (TNF-α), interleukin-2 (IL-2), interleukin-4 (IL-4), interleukin-10 (IL-10), their plasma levels being associated with disease severity, leading to a so-called “*cytokine storm*”, thus explaining some young adults’ disease severity (29).

Immune system hyper-reaction was also described in the 1918–1919 influenza pandemic, which was the first known pandemic to report an excess risk of death among individuals 25–35 years old (9). Few studies proved that influenza aggravated pulmonary status of individuals with TB, so that latent TB could become active, a closed cavity might open, various lesions might progress leading to further deterioration of pulmonary function (9).

Cytokines have an important role in host resistance to TB infection, being first demonstrated in murine infection models (21) and later validated by severe mycobacterial disease findings in patients with mutations in the IFN-*γ* and IL-12 signalling pathways and in rheumatoid arthritis or Crohn’s disease patients treated with TNF-α blockade (21, 30).

Since the SARS-CoV-2 is a newly discovered pathogen (first infection being reported in December 2019) (29), little data about the coinfection with MTB could be found (especially considering the long incubation period of MTB from exposure to developing the disease, often with a slow onset) (31, 32). Still, the existent studies show that TB status plays a major role in a rapid development of severe acute respiratory syndrome in SARS-CoV-2 coinfection considering the cases described in China (14) and India (33).

One should keep in mind that the existence of underlying conditions, autoimmune diseases, poor hygiene, and overcrowding are all known as risk factors for developing one, another or both diseases (29, 31), as it was suggested in a paper developing a model of pathogen dissemination in the outpatient clinic, that populations with high risk of contracting influenza or SARS might also have higher prevalence of MTB (34).

#### Keypoints

1. Cytokines seem to play an important role in both COVID-19 and TB, their plasma level being associated with disease’s severity.
2. Immune system hyper-reaction could explain a poorer outcome in people 25–35 years old.
3. Although there is limited data on MTB and COVID-19 coinfection, one could reasonable presume that their coexistence might have a more severe evolution for the patient.

### 4.3. (Proven or presumed) clinical and paraclinical impacts of vaccination

One of the most effective ways to prevent diseases caused by pathogens like bacteria or viruses proved to be vaccination (35). Since the first discovery of SARS, extensive research was made for finding a vaccine to prevent the onset of the disease (36). Different vaccine types were tested: inactivated or live-attenuated virus, DNA-based vaccines, recombinant proteins, virus-like particles, viral vectors with some promising efficiency, but with neither being finally approved for use (36, 37). Recent data suggest that SARS-CoV-2 genome is up to 80% similar to SARS-CoV-1 and up to 50% similar to MERS-CoV (38), so previous studies on protective immune responses against SARS-CoV-1 or MERS-CoV may aid vaccine development for SARS-CoV-2 (39, 40). Considering that to date there is no approved vaccine neither for SARS-CoV-1 nor for MERS-CoV, other options are considered, such as the vaccine used for TB prevention (41).

Since 1921 a vaccine is used widely for TB prevention - a live attenuated strain of the bovine tubercle bacillus named bacillus Calmette-Guerin (BCG) (42, 43). In 1927 was observed that BCG-vaccinated new-borns had a three times lower mortality rate in their first year of life than the unvaccinated ones (44). Later was noted a decrease in infectious morbidities, protecting both mice (against secondary fungal or parasitic infections with *Candida albicans* or *Schistosoma mansoni* through tissue macrophages activation) (42) and infants (against acute lower respiratory infections). Thus, the risk of acute lower respiratory infections in BCG-vaccinated infants seemed to be 37% lower than in unvaccinated controls (42, 45).

There is data suggesting that BCG vaccination of adults could increase the capacity of producing pro-inflammatory cytokines such as Il-1β and IL-6 which leads to non-specific protection against unrelated pathogens like *Staphylococcus aureus* or *Candida albicans* (46).

Considering these facts, BCG vaccine is contemplated as a potential candidate against respiratory viruses (41). Moreover, Muldron Childrens Research Institute from Australia already announced a phase III randomised controlled trial which will determine if health-care workers’ BCG vaccination will have any impact on SARS-CoV-2 infection (BCG Vaccination to Protect Healthcare Workers Against COVID-19, BRACE, NCT04327206) but more time is needed to establish its’ supposed efficiency.

It is noted that SARS-CoV-2 envelope spike (S) protein has a decisive role for determining host tropism and transmission capacity (38) and T cell epitopes-based peptide derived from S proteins that map to SARS-CoV-2 proteins (39) and subunit vaccines based on S protein are also considered for preventing SARS-CoV-2 infection (39, 47–50).

Novel methods are emerging such as reverse vaccinology that refers to the process of constructing vaccines by detecting viral antigens through genomic analysis using bioinformatics tools. Reverse vaccinology has successfully been applied in the fight against Zika virus or Chikungunya virus (51). One study proposed reverse vaccinology and immunoinformatics methods to design potential subunit vaccines against SARS-CoV-2 using the highly antigenic viral proteins and epitopes. Suggested vaccine constructs appeared to confer good immunogenic response through various computational studies. Three vaccine constructs were designed and the best one was selected through molecular docking study (51). Another study proposes a specific synthetic vaccine epitope and peptidomimetic agent, identified through bioinformatics methods (52).

Currently, there are 15 potential vaccine candidates for SARS-CoV-2 in the pipeline globally developed using various technologies (messenger RNA, synthetic DNA, synthetic and modified virus-like particles) (53–55).

#### Keypoints

1. SARS-CoV-2 genome is up to 80% similar to SARS-CoV-1 and 50% similar to MERS-CoV
2. No SARS vaccine was approved for clinical use (in 18 years of research).
3. Ongoing trials on SARS-CoV-2 vaccine are on the highest interest.

### 4.4. Diagnostic errors in the context of COVID-19 & TB coexistence (or, how does one condition influence diagnosis of the other one?)

TB and COVID-19 are mainly respiratory diseases that affect primarily the lungs; however, the onset of TB is often slow compared to COVID-19 which seems to develop in a few days from exposure (4, 23). Given the clinical and imagistic similarities such as cough, fever or shortness of breath and various radiological pulmonary lesions (4, 23), accurate diagnostic tests should be made available in order not to overlook one condition in favour of the other one.

Tuberculin skin test (TST), and with a greater sensibility and specificity the interferon-gamma release assays (IGRAs), are widely used for TB screening (56). Given their results are influenced by the host’s immune response after MTB (or BCG) exposure (57), there is a gap for diagnostic errors in individuals with an impaired immune system, such as in a concurrent severe infection (58, 59). Increased age, low peripheral lymphocyte count, high body mass index and immunosuppressive therapies were also associated with false negative results (59) that could lead to missing TB diagnose. Moreover, an excess of inflammatory markers could affect IGRAs sensitivity and the high value of C-reactive protein (CRP) might be a confounder for false negative results (60).

It has been observed that high CRP and low peripheral lymphocyte counts could occur within a few days of exposure to SARS-CoV-2 (61). Therefore, this observation may lead to the possibility that a patient with latent TB or TB sequel may have a false negative IGRA.

As SARS-CoV-2 has not been identified since few months in humans, there is no specific treatment (13). Given the growing number of reported cases, it is important that suspected patients be diagnosed as quickly as possible to isolate and limit further transmission (13). Conventional methods such as assays for detection of viral antigens or antiviral antibodies and newer methods of diagnosis as multiplex nucleic acid amplification have been developed and used clinically (13).

With the urge of identifying the radiological features of SARS-COV-2 infection, with the community transmission present in most countries, and with its non-specific clinical onset (fever, dry cough, dyspnoea and radiological findings of bilateral infiltrates and even pleural effusion and cavitation) (62) doctors may be facing either a difficult differential diagnostic, either not consider tuberculosis at all.

Considering the sudden onset of the SARS-CoV-2 pandemic, countries struggled to quickly find a possible treatment to prevent respiratory failures and deaths especially among patients with respiratory comorbidities. In addition, since its fatal dinamics, there is no time to carry out new drug development in the traditional manner. Therefore, screening for already available drugs (for any activity against SARS-CoV-2) (13) is usually preferred at the first instance. It seems that an antiviral used for HIV infection, composed of two protease inhibitors (lopinavir and ritonavir), would have a therapeutic effect on coronavirus infections. It seems to have entered as a recommendation in the treatment of the COVID-19 in a short time (13). Other compounds such as redexivir, favivir, ribavirin, nitrazine, chloroquine / hydroxychloroquine are being evaluated (13, 61). Chloroquine and hydroxychloroquine have been shown to shorten the duration of SARS-CoV-2 viremia by reducing the viral load (61). However, hydroxychloroquine has also been associated with a higher risk of nontuberculous mycobacterial (NTM) infection in rheumatoid arthritis patients (63).

#### Keypoints

1. Coinfection of TB and SARS-COV-2 may be difficult to diagnose.
2. SARS-COV-2 infection may mask the clinical and radiological active TB.
3. Patients receiving the proposed treatment for COVID19 may be at risk for the infection with NTM.

## 5. Conclusions

Because viral respiratory infections and TB impede the host's immune responses, their harmful synergism can be assumed to contribute to more severe clinical evolution. Coinfection most likely affects both sides of these patients: rapid development of severe acute respiratory syndrome through cytokine-mediated immune response, as well as increased risk of tuberculosis reactivation. As a lesson from previous outbreaks, hospital treatment for patients with tuberculosis should be limited to severe cases, to prevent the spread of SARS-CoV-2 in TB cases. Despite the rapidly increasing number of cases, the data needed to predict the impact of the COVID-19 pandemic on patients with latent TB and TB sequelae and to guide management in this particular context still lies ahead.

## Data Availability

Data used to support the findings of this study are available from the corresponding author upon request.

